# Hypertension detection, treatment and control in young adults in Australia

**DOI:** 10.64898/2026.01.27.26345008

**Authors:** Ritu Trivedi, Niamh Chapman, Michael O Falster, Andrew E. Moran, Aletta E Schutte, Bin Zhou, Dean S Picone

## Abstract

**Background:** Young adult hypertension increases risk of future cardiovascular disease (CVD) but is often overlooked. We aimed to identify opportunities to reduce blood pressure (BP) among younger adults by determining the drivers of hypertension detection, treatment and control in young adults in Australia.

**Methods:** Cross-sectional analysis of adults aged 18–39 years using nationally representative 2017-18 Australian National Health Survey data. Descriptive analyses were performed to determine hypertension prevalence, detection, treatment and control by sex, health services interaction, geographical location and socioeconomic status, with subgroup analyses among those with co-existing cardiometabolic risk factors. Hypertension was defined as self-reported diagnosis, treatment, and/or measured BP≥140/90mmHg. Detection was defined as self-reported hypertension, treatment as taking ≥1 antihypertensive class, and control as BP<140/90mmHg.

**Results:** The survey sample (n=5,217) was representative of N=7,533,625 adults aged 18–39 years (49.9% women). Hypertension prevalence was 11.2% (95% CI: 11.1% to 11.2%). Among those with hypertension, detection was 13.5%, treatment was 6.5% and control was 4.3%. These rates were influenced by factors related to healthcare access, for example, young adults who had a recent BP check and resided in major cities had higher rates than their counterparts.

**Conclusions:** One in ten young Australians adult have hypertension, yet only 13.5% are detected. Differences in detection, treatment and control rates were partly explained by factors related to healthcare access. There is a need to better detect and manage high BP in young adults, potentially by developing tailored approaches that are specific to different contexts and ultimately supporting prevention of CVD in later life.

## INTRODUCTION

Hypertension is the leading modifiable risk factor for cardiovascular disease (CVD) affecting 1.3 billion people worldwide^1^, including 6.8 million Australians (one-third of the adult population)^2^. Whilst older adults account for the majority of people with hypertension, many younger adults also have hypertension^3^. High blood pressure (BP) in this group is important because vascular and end-organ damage from lifetime chronically uncontrolled BP is cumulative, and increases the likelihood of developing clinical CVD later in life^4-7^. Young adults with BP greater than 130/80 mmHg, compared to those within normal ranges, are at significantly higher risk for subsequent cardiovascular events^4,7^. Even people with elevated BP (systolic BP 120 to 129 mmHg) before the age of 40 years are at increased risk of CVD compared to young adults with normal BPs^5^ and, those who develop high BP at an older age, due to the cumulative exposure to high BP over time^6^.

In Australia, guidelines recommend initiation of treatment of hypertension for primary prevention of CVD based on 5-year absolute CVD risk score^8^. Pharmacological treatment based solely on BP values alone, regardless of CVD risk score, is recommended when BP is above 160/100 mmHg^8^. However, most young adults fall into the ‘low’ CVD risk category due to younger age, and therefore, may miss out on the opportunity to modify lifestyle or receive pharmacological treatment of high BP. Additionally, younger adults are less engaged in the healthcare system and therefore may have fewer opportunities for hypertension detection, thus remaining unaware of high BP^9^. Few studies specifically describe trends in hypertension detection, treatment and control in young adults, especially in Australia^9,10^. Due to significant later-life CVD-related morbidity and mortality caused by young adult hypertension, understanding this data is necessary to develop targeted preventative health intervention strategies. This study aimed to quantify hypertension prevalence, and its detection, treatment and control in young Australian adults.

## METHODS

### Overview

A cross-sectional analysis of hypertension prevalence, detection, treatment and control of young Australian adults (aged 18–39 years) was undertaken. Nationally representative population characteristics were obtained from the Australian National Health Survey 2017-18 publicly available aggregated data^2^. This survey collected comprehensive health-related data from Australians living in private dwellings across urban and rural areas. The survey used a stratified multistage sample design to select approximately 21,300 individuals from 16,400 households across all major cities, inner regions and rural areas in Australia. Within each household, one adult (aged 18 years and over) and one child were randomly selected for participation. Trained interviewers conducted face-to-face interviews to gather self-reported data on health conditions, risk factors, medications and health service usage and measured the individual’s BP. More details are available in the online Supporting Information.

### Blood pressure measurements

Consenting adults had BP measured in a seated position using an automated monitor with at least two readings and the second was used for analysis. If the difference between the two readings was greater than 10 mmHg a third reading was taken and the second and third were averaged for analysis unless the third differed by 20 mmHg or more, in which case the readings were considered invalid. Unavailable data was imputed using the ‘hot decking’ method to match population characteristics, including age, sex, location and other self-report variables, between missing data and measured data^2^.

### Hypertension prevalence

Hypertension was defined as people who self-reported receiving a diagnosis from a clinician, and/or were taking antihypertensive medications (at least one medicine grouped as either a diuretic, beta blocker, calcium channel blocker or agent acting on the renin-angiotensin system), and/or had measured BP ≥140/90 mmHg, similar to reporting in other global studies^11,12^.

### Hypertension detection, treatment and control

Detection was defined as those with hypertension and self-reported having hypertensive diseases (including hypertension) on the survey. Treatment included those that reported taking BP lowering medications grouped by the following classes (based on WHO’s Anatomical Therapeutic Chemical medication codes): diuretics, beta blocking agents, calcium channel blockers, agents acting on the renin-angiotensin system. Control was defined as measured BP <140/90 mmHg.

### Sociodemographic and clinical characteristics

Health service interaction was assessed based on self-reported recent BP checks, categorised as checked within the last two years, and not checked or unknown if checked within the last two years. Geographical location was classified using the Australian Statistical Geography Standard (ASGS) Remoteness Structure into major cities, inner regional, and rural areas (including outer regional, remote, and very remote). Socioeconomic status was assessed using the Socio-Economic Indexes for Areas (SEIFA) Index of Relative Socio-Economic Disadvantage, a composite measure that ranks areas based on variables such as income, education, employment, and housing characteristics, grouped into quintiles (where Q1 represents the most disadvantaged and Q5 represents the least disadvantaged). Current additional cardiometabolic risk factors included high cholesterol, based on self-reported diagnosis by a health professional and obesity, defined as a body mass index (BMI) ≥ 30 kg/m^2^ calculated from measured height and weight.

### Statistical analysis

Descriptive analyses on demographics, clinical characteristics and BP are presented as weighted means ± SD or weighted proportions (%) of the survey sample. Analyses were conducted using R and SPSS, and were stratified by sex, recent BP checks, geographic location and socioeconomic status. Separate subgroup analyses were conducted among people with cardiometabolic risk factors of high cholesterol and obesity. A sensitivity analysis was conducted to reclassify those only taking beta-blocker medications, and who did not self-report or have measured high BP, as not having hypertension because this medication may be used for other purposes beyond BP in young adults.

## RESULTS

### Participant characteristics

The survey sample included a total of 5,217 young adults, which was representative of 7,533,625 adults aged 18 to 39 years (49.9% women) (Table 1). Just over three-quarters of participants resided in major cities, and about one-third held a bachelor or postgraduate degree. The overall average BP was 114±14 mmHg and 56% of people were overweight or obese. A total of 82% of people reported having their BP checked in the prior two years and 1.5% of participants reported receiving antihypertensive treatment with at least one medication class (Table 1).

**Table 1.** Population characteristics of 18–39-year-olds Australian adults.

|  |  | Female | Male | Total |
| --- | --- | --- | --- | --- |
| **Weighted population total, N** |  | 3,755,435 | 3,778,190 | 7,533,625 |
| **Unweighted survey sample total, n** |  | 2,775 | 2,442 | 5,217 |
| **Geographical rurality** |  |  |  |  |
|  | Major cities | 78.0% | 78.1% | 78.1% |
|  | Inner regional | 14.0% | 14.5% | 14.3% |
|  | Rural (outer regional, remote, very remote) | 8.0% | 7.4% | 7.7% |
| **Highest education attainment** |  |  |  |  |
|  | Below Year 12 | 12.4% | 15.3% | 13.9% |
|  | Year 12 or equivalent | 21.0% | 22.9% | 22.0% |
|  | Advanced diploma/ Diploma/ Certificate | 27.3% | 33.6% | 30.4% |
|  | Bachelor/ Postgraduate degree | 39.4% | 28.2% | 33.7% |
| **Employment status** |  |  |  |  |
|  | Employed | 75.5% | 85.5% | 80.5% |
|  | Unemployed | 4.5% | 5.0% | 4.8% |
|  | Not in the labour force | 20.1% | 9.4% | 14.7% |
| **Index of Relative Socio-Economic Disadvantage** |  |  |  |  |
|  | 1<sup>st</sup> quintile = most disadvantaged | 18.1% | 17.2% | 17.6% |
|  | 2<sup>nd</sup> quintile | 18.8% | 20.8% | 19.8% |
|  | 3<sup>rd</sup> quintile | 23.4% | 22.1% | 22.7% |
|  | 4<sup>th</sup> quintile | 22.1% | 19.6% | 20.8% |
|  | 5<sup>th</sup> quintile | 17.6% | 20.3% | 19.0% |
| **Body mass index\*** |  |  |  |  |
|  | Healthy | 50.0% | 33.8% | 41.9% |
|  | Overweight | 26.6% | 40.8% | 33.7% |
|  | Obese | 20.7% | 23.8% | 22.3% |
|  | Underweight | 2.7% | 1.5% | 2.1% |
| **Measured blood pressure** |  |  |  |  |
|  | Systolic, mean ± SD, mmHg | 109 ± 12 | 120 ± 14. | 114 ± 14 |
|  | Diastolic, mean ± SD, mmHg | 73 ± 10 | 75 ± 11 | 74 ± 10 |
|  | Classification<sup>†</sup> |  |  |  |
|  | Optimal | 70.4% | 41.1% | 55.7% |
|  | Normal | 15.8% | 31.2% | 23.5% |
|  | High-normal | 7.0% | 15.9% | 11.5% |
|  | Hypertension |  |  |  |
|  | Grade 1 (mild) | 5.6% | 9.9% | 7.7% |
|  | Grade 2 (moderate) or higher | 1.2% | 2.0% | 1.6% |
|  | Hypertension subtype |  |  |  |
|  | Isolated systolic | 0.6% | 5.2% | 2.9% |
|  | Isolated diastolic | 4.9% | 3.9% | 4.4% |
|  | Both systolic and diastolic | 1.3% | 2.8% | 2.0% |
| **Recent blood pressure checks** |  |  |  |  |
|  | Blood pressure checked in last 2 years | 88.7% | 75.8% | 82.2% |
|  | Blood pressure not checked in last 2 years | 10.0% | 22.3% | 16.1% |
|  | Unsure if blood pressure was checked in last 2 years | 1.4% | 1.9% | 1.6% |
| **Medical history<sup>‡</sup>** |  |  |  |  |
|  | Hypertension<sup>§</sup> | 8.4% | 13.9% | 11.2% |
|  | High cholesterol | 0.7% | 0.9% | 0.8% |
|  | Diabetes type 1 | 0.3% | 0.8% | 0.5% |
|  | Diabetes type 2 | 0.3% | 0.5% | 0.4% |
|  | Stroke | 0.2% | 0.2% | 0.2% |
|  | Angina | 0.1% | 0.2% | 0.1% |
|  | Heart attack | 0.0% | 0.1% | 0.0% |
|  | Heart failure | 0.1% | 0.0% | 0.1% |
|  | Other heart disease | 0.0% | 0.2% | 0.1% |
|  | Other ischemic diseases | 0.2% | 0.1% | 0.1% |
| **Treatment** |  |  |  |  |
|  | Antihypertensive classes |  |  |  |
|  | Renin-angiotensin system agents | 0.7% | 1.0% | 0.9% |
|  | Diuretics | 0.3% | 0.0% | 0.2% |
|  | Calcium channel blockers | 0.2% | 0.0% | 0.1% |
|  | Beta blocking agents | 0.7% | 0.7% | 0.7% |
|  | Antihypertensive, number of classes |  |  |  |
|  | One | 1.4% | 1.3% | 1.3% |
|  | Two or more | 0.2% | 0.3% | 0.2% |

*Footnote:* All reported percentages (%) are weighted. \*Body mass index categories: <18.5 kgm<sup>-2</sup> (underweight), ≥18.5 to <25.0 kgm<sup>-2</sup> (healthy), ≥25.0 to <30.0 kgm<sup>-2</sup> (overweight) and ≥30.0 kgm<sup>-2</sup> (obese). <sup>†</sup>measured blood pressure was categorised according to thresholds in the most recent Australian hypertension guidelines from 2016: optimal (<120/80 mmHg), normal (120/80 to 129/84 mmHg), high-normal (130/85 to 139/89 mmHg), grade 1 (mild) hypertension (140/90 to 159/99 mmHg), grade 2 (moderate) hypertension or higher (≥160/100 mmHg). <sup>‡</sup>conditions were mostly self-reported, except for <sup>§</sup>hypertension which was defined as people who had measured high blood pressure (≥140/90 mmHg), or normal measured blood pressure and were taking antihypertensive medications.

### Hypertension prevalence

#### Overall and sex-specific

The prevalence of hypertension in young adults was 11.2% (95% CI: 11.1% to 11.2%) with higher prevalence in men compared to women (men: 13.9%, 95% CI: 13.87% to 13.94%; women: 8.4%, 95% CI: 8.35% to 8.41%). Classification of hypertension of the entire cohort and among young adults with hypertension is available in Table 1 and Supporting Information Table S1, respectively.

#### Recent BP measurement, geographical location and socioeconomic status

The prevalence of hypertension in young adults across various subgroups was similar: recent BP measurement (checked within last two years: 11.3%, not checked or not known if checked in last two years: 10.4%), geographical locations (major cities: 10.6%, inner regional: 12.6% and rural areas: 14.3%) and SEIFA quintiles (Q1: 12.6%, Q2: 12.0%, Q3: 10.7%, Q4: 10.9%, Q5: 9.9%) (Supporting Information Table S2).

#### Additional cardiometabolic risk factors

The prevalence of hypertension in young adults with high cholesterol (51.0%) and obesity (22.1%) was higher than in the general population (11.2%) (Supporting Information Table S2).

### Hypertension detection, treatment and control

#### Overall, including sex specific

Among young adults with hypertension, 13.5% were detected, 6.5% were treated and 4.3% were controlled to <140/90 mmHg (Supporting Information Figure S1). Detection (16.2% vs. 11.9%), treatment (9.6% vs. 4.7%) and control (5.2% vs. 3.7%) was better in women compared with men (Figure 1).

**Figure 1.**
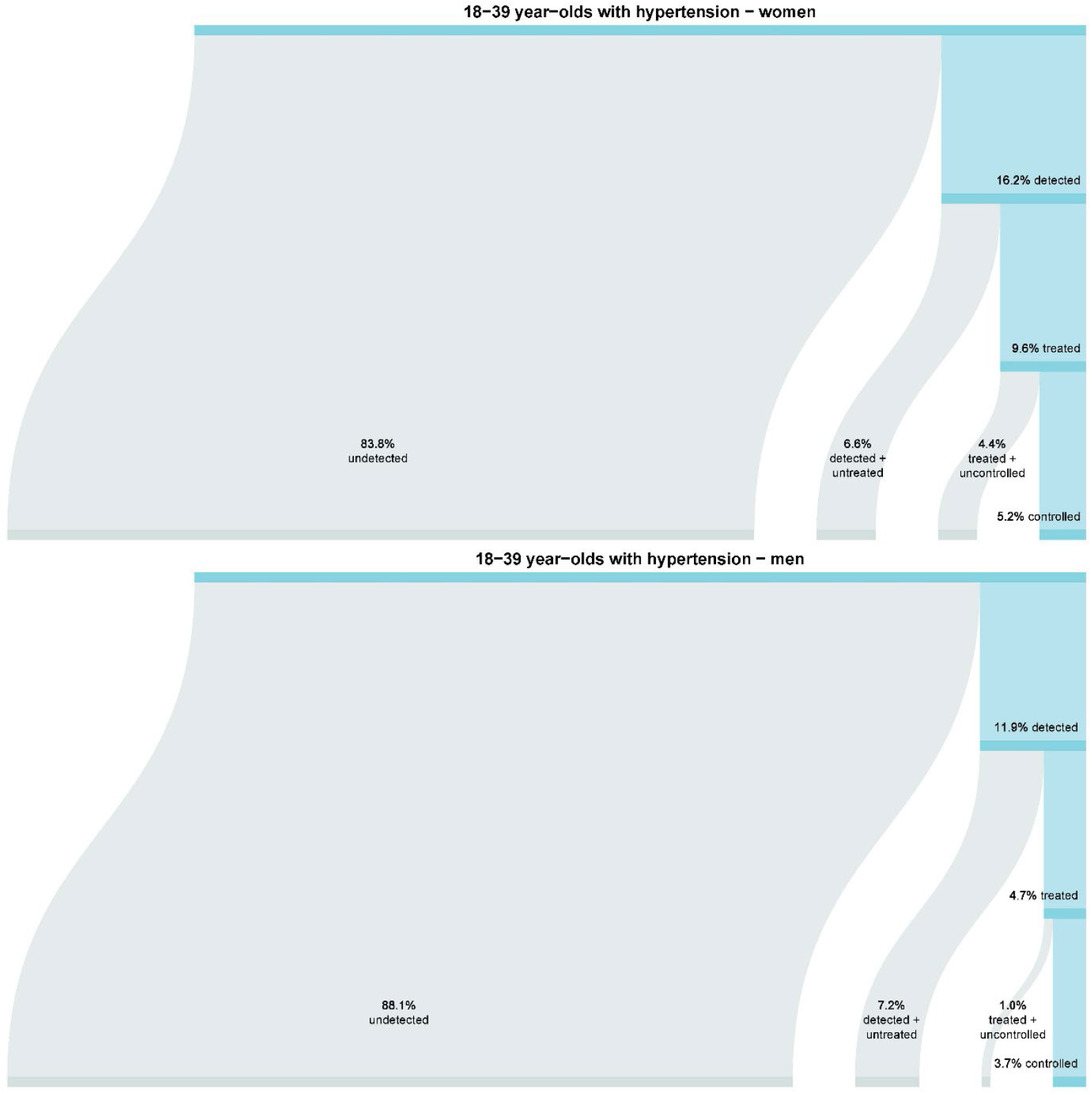
Detection, treatment and control of hypertension in (a) women and (b) men aged 18 to 39 years.

#### Recent BP measurement, geographic location and socioeconomic status

Detection, treatment and control was higher among those that had their BP checked in the last 2 years (15.8%, 7.8% and 5.1%, respectively) compared to those that did not where only 2.2% had hypertension detected with 0% treated or controlled (Figure 2). Hypertension detection, treatment and control were all relatively similar in major cities and inner regional areas and slightly higher than the rates of detection, treatment and control in rural areas (Figure 3). Hypertension detection and treatment were similar in the highest and lowest socio-economic subgroups (Supporting Information Figure S2). Hypertension control was slightly better in the highest socio-economic subgroup compared with the other subgroups (Supporting Information Figure S2).

**Figure 2.**
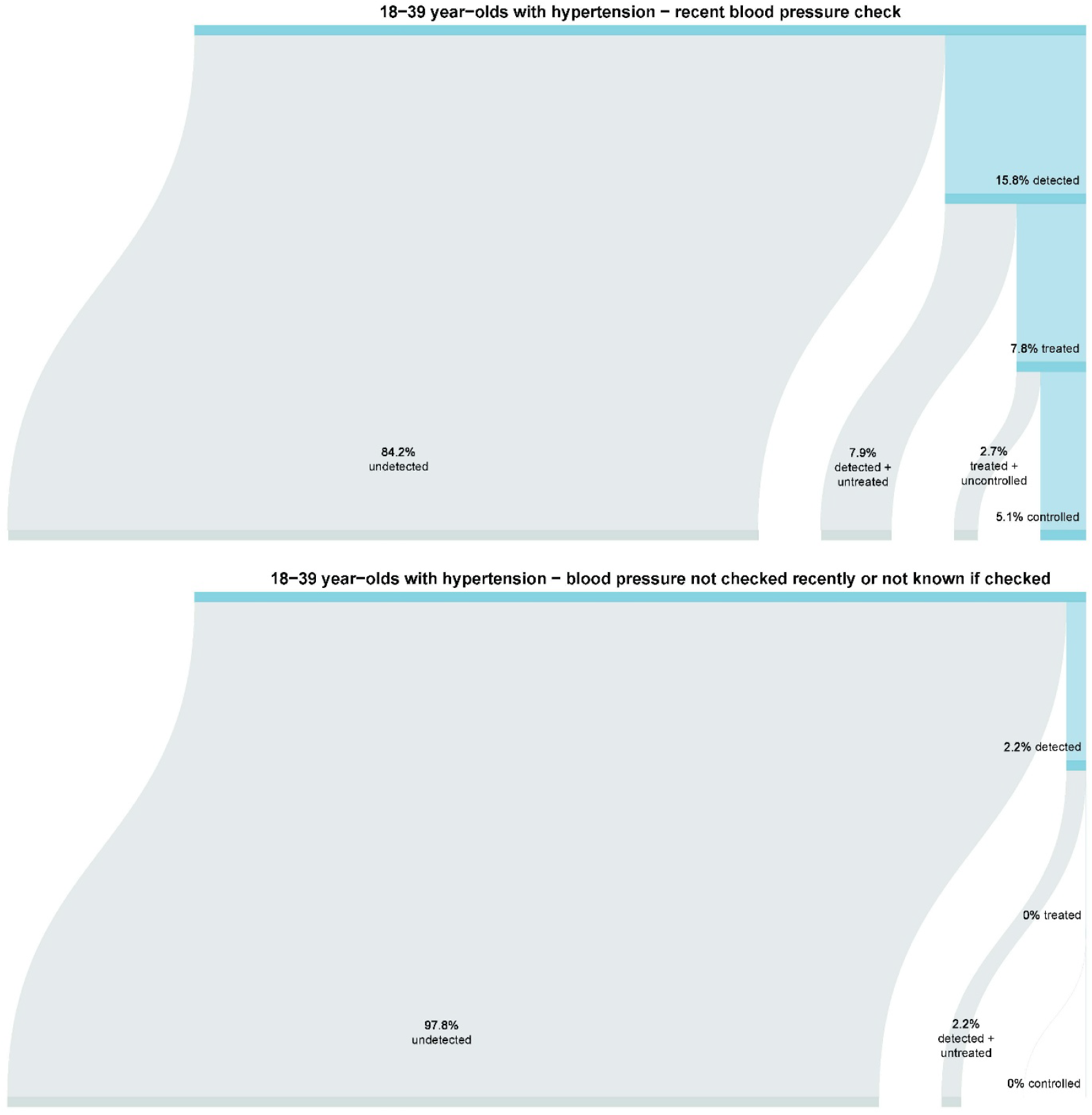
Detection, treatment and control of hypertension in those aged 18 to 39 years by whether blood pressure was recently (a) checked, (b) not checked or not known if checked

**Figure 3.**
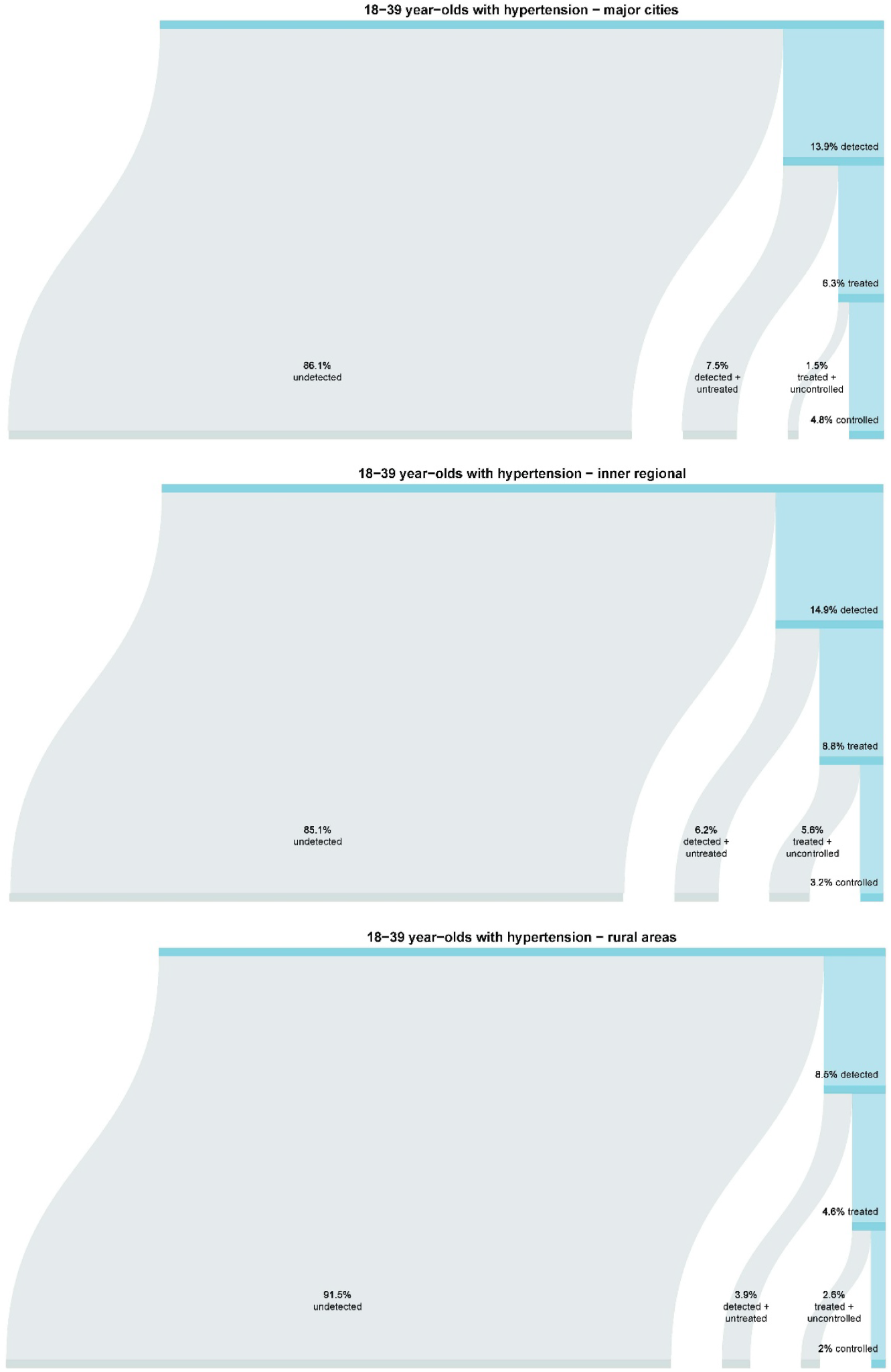
Detection, treatment and control of hypertension in those aged 18 to 39 years by geographical location: (a) major cities, (b) inner regional and (c) rural areas (outer regional or remote areas).

#### Additional cardiometabolic risk factors

In the subgroup of young adults with high cholesterol, rates of detection (86.8% vs. 13.5%), treatment (64.2% vs. 6.5%) and control (34.2% vs. 4.3%) were all higher than analyses among the general population (Figure 4). In the subgroup of young adults with obesity, hypertension detection and treatment were similar to the general population, but control (1.3% vs. 6.4%) was poorer (Supporting Information Figure S3).

**Figure 4.**
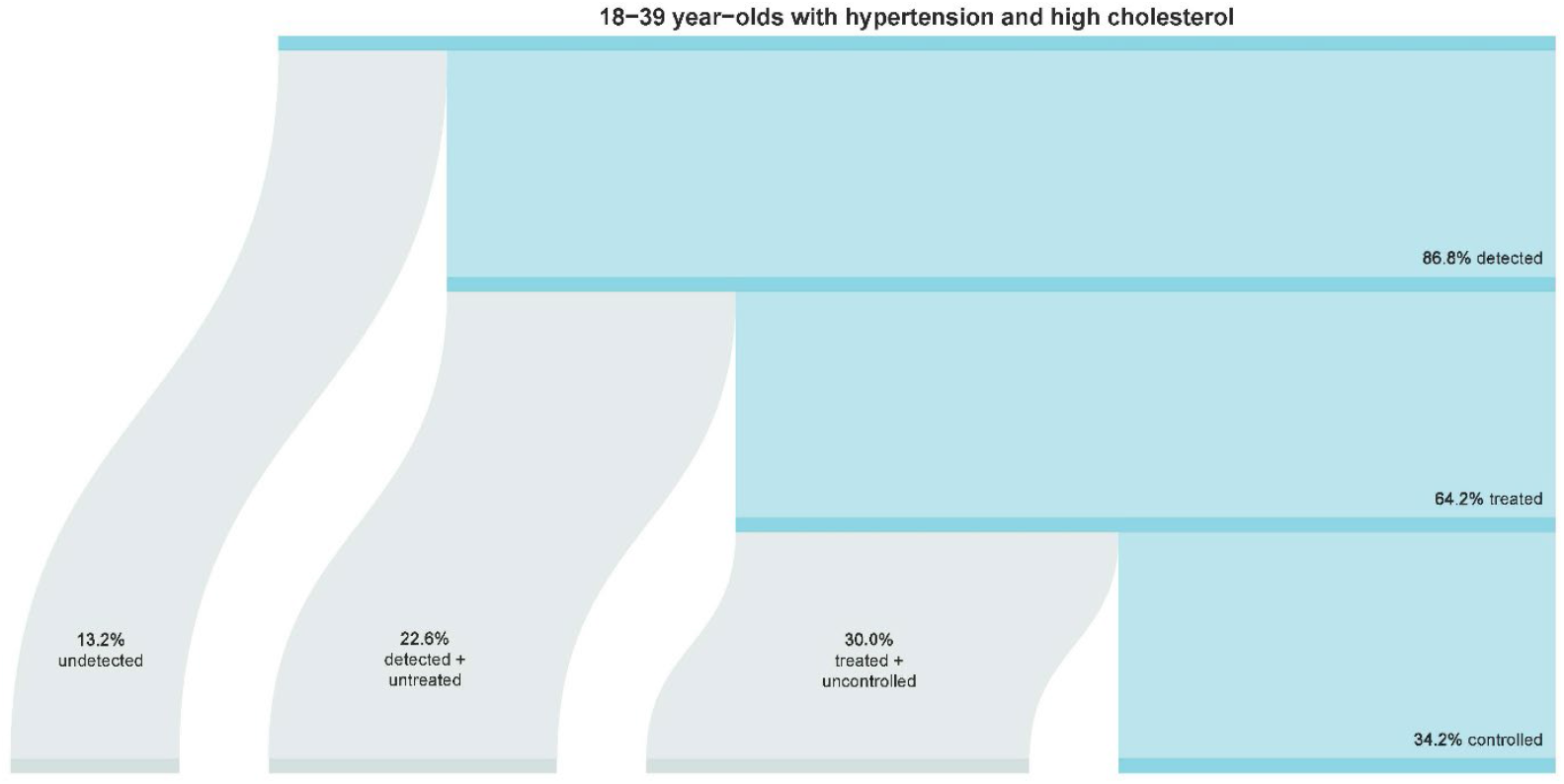
Detection, treatment and control of hypertension in those aged 18 to 39 years with high cholesterol.

### Sensitivity analysis

When those who were taking only beta-blockers and did not meet other hypertension definition criteria (ie, self-reported or had high measured BP) were reclassified as not having hypertension, similar prevalence (10.8%), detection (13.9%), treatment (6.7%) and control (4.4%) rates were observed as in the main analysis (Supporting Information Figure S4).

## DISCUSSION

This analysis of nationally representative data from Australia revealed that one in ten young adults had hypertension, yet only 13.5% of young adults living with hypertension had their condition previously detected. There were several other notable novel findings related to healthcare access. First, among the 83% of young adults with hypertension that reported their BP was checked in the previous two years only 16% had hypertension detected. Second, detection of hypertension was higher in major cities and inner regional areas. Third, among people with high cholesterol and hypertension, there was substantially better detection, treatment and control rates than in the general young adult population. Overall, the most pressing concerns was the low detection of hypertension, particularly among young men and low rates of treatment initiation. These findings underscore the need for preventative health strategies to support identification, confirmation and management of high BP to lower risk of future CVD in young adults in Australia.

Approximately one in ten young adults in Australia have hypertension^9^, placing them at substantially high risk for CVD, dementia, and premature mortality compared to those with normal BP^4,5,13,14^. Young adult hypertension is likely to become more prevalent in the upcoming years. This is because the global prevalence of hypertension in children and adolescents doubled between 2000 and 2020^15^ and that BP tracks from childhood to adulthood^16^. Rising prevalence of earlier onset hypertension may expose more young people to increased risk of CVD. Indeed, a meta-analysis, totalling 4.5 million people aged 18 to 45 years, showed continuous and graded associations between categorical BP increments and increased risk of cardiovascular events, coronary heart disease, stroke and all-cause mortality^13^. There is also emerging evidence on the association between mid-life high BP and future dementia^17^. The strong association between elevated BP in young adults and future adverse health outcomes shows the need to raise awareness of the importance of maintaining normal BP levels, establish effective pathways for detection and confirmation, and implement preventative health strategies that are acceptable to young adults and appropriate in different contexts to lower BP.

In the present study 6.5% of young Australian adults with hypertension were treated, contrasting the United States where about 50% are treated^9^. A recent meta-analysis of Australian studies found a similarly low hypertension treatment rate among young adults^3^. These data suggest that Australia does not have well defined, standardised pathways to support detection, treatment and control of hypertension for young adults. The reasons for low treatment rates among young adults are highly complex and multifactorial. First, the perception of hypertension being an “older person’s disease” may negatively impact patient motivation to seek out healthcare services^18,19^. However, 86% of young adults in this study had a BP check in the previous two years, indicating that most people in this age range are engaged in some type of health check. Second, clinical inertia may occur due to lack of randomised trial evidence on treatment in young adults or long-term outcome data, along with concerns about the risk-to-benefit ratio of treatment in young adults^20,21^. Alternative evidence sources are beginning to emerge, with a recent pharmacoepidemiology analysis showing potential benefits of antihypertensive treatment in young adults by demonstrating lower risk of CVD hospitalisations among patients adherent to medication compared with those who were not^22^. Third, in Australia, guidelines for CVD risk prevention recommend absolute CVD risk assessment to support clinical decisions and that only when BP is ≥160/100 mmHg should BP lowering treatment be initiated irrespective of CVD risk. For younger adults with hypertension, risk of CVD in the next 5-10 years is often low, especially in people with few or no additional risk factors, which may mean that there is no indication for action to be taken based on the calculated absolute CVD risk. In contrast to this approach, in the 2025 update of the American Heart Association/American College of Cardiology hypertension guidelines, initiation of BP lowering medications in all adults whose systolic BP remains ≥ 130 or diastolic BP ≥ 80 mmHg after 3-6 months of lifestyle intervention, even if their absolute CVD risk is not in the high category^23^. If similar changes to recommendations were to be made in Australia, this would increase costs to patients and health services but may result in substantial cost savings due to improved global domestic product, quality adjusted life years and decreased CVD events in future years^24^. However, risk communication strategies will be important to present the potential benefits of medicines alongside the risk of long-term exposure to high BP levels to support shared decision making in this cohort.

Tailored strategies that engage with young adults in ways that reach beyond traditional healthcare visits to correctly identify hypertension could be impactful in improving low hypertension detection rates. The present study showed that disparities in detection, treatment and control outcomes were evident in young adults with lower healthcare access compared to their counterparts, such as people who had not had a recent BP check and residing in rural areas. To address issues related to access, workplace screening^25^ and community-based care ^26^ could offer an alternative to traditional healthcare settings for engaging with young adults. Other alternatives that offer promise for CVD prevention include the growing market of digital health technologies, which both are particularly popular among younger people and support healthcare outside of traditional settings^27^. These technologies can range from motivational messages or applications for healthy lifestyle behaviours and medication adherence nudges through to devices for self-monitoring BP, including emerging wearable cuffless devices^27-29^. However, the clinical effectiveness of some of these technologies and their implementation and integration into clinical practice is yet to be established^28^. Newer models of care that can accommodate increased demands amid anticipated healthcare workforce shortages are a necessary step towards improving cardiovascular health.

### Strengths and limitations

A major strength of this study was the use of data that was nationally representative of 7.5 million young adults 18-39 years old^2^. However, some limitations need to be considered. First, the most recent aggregated survey data from the Australian National Health Survey was only available from years 2017-18. We do not anticipate any major differences in results when newer data becomes available based on the summary statistics of the 2020-21 National Health Survey and a recent meta-analysis pooling studies conducted in Australia between 1980 and 2018^9^. These both reported similar hypertension prevalence by age as in this current study. Moreover, Australian guidelines regarding absolute CVD risk management have remained consistent. Our study presented more in-depth results on hypertension in young adults than the aggregate numbers in the aforementioned reports. Second, the analyses were based on BP readings that were measured as part of the survey interview process, and about 32% of participants did not have their BP measured. The missing data was imputed based on population demographics per standard statistical approaches^2^. Third, medication data which informed the treatment rates were based on self-reported medications taken in the past two weeks, which were then categorised based on the ATC medication codes. If either diuretic, calcium channel blockers, renin-angiotensin agents and beta-blocker categories were indicated then this was considered as taking ‘antihypertensive treatment’, however, there may be other reasons people were taking these medications, especially beta-blockers. A sensitivity analysis to declassify hypertension in people who reported taking only beta-blockers, had not self-reported having hypertension and had a normal measured BP reading revealed similar results in prevalence, detection, treatment and control rates.

### Conclusion

In conclusion, approximately 1 in 10 young adults in Australia have hypertension. Rates of detection, treatment and control are low among Australian young adults, particularly those with reduced access to healthcare. These results reiterate the need for tailored preventative health approaches to improve detection, diagnosis and overall management of hypertension in young Australian adults. Specifically, clear clinical guidance for young adults is needed to tackle inertia for diagnosis and treatment initiation, which will likely need to be tailored for specific contexts.

## Data Availability

As we are not the data custodians, we are not authorised to make the data available. With appropriate approvals, the data may be accessed through the Australian Bureau of Statistics.

